# Increased risk of infection with SARS-CoV-2 Beta, Gamma, and Delta variant compared to Alpha variant in vaccinated individuals

**DOI:** 10.1101/2021.11.24.21266735

**Authors:** Stijn P. Andeweg, Harry Vennema, Irene Veldhuijzen, Naomi Smorenburg, Dennis Schmitz, Florian Zwagemaker, SeqNeth Molecular surveillance group, RIVM COVID-19 Molecular epidemiology group, Arianne B. van Gageldonk-Lafeber, Susan J.M. Hahné, Chantal Reusken, Mirjam J. Knol, Dirk Eggink

## Abstract

The extent to which severe acute respiratory syndrome coronavirus 2 (SARS-CoV-2) variants of concern (VOC) break through infection- or vaccine-induced immunity is not well understood. Here, we analyze 28,578 sequenced SARS-CoV-2 samples from individuals with known immune status obtained through national community testing in the Netherlands from March to August 2021. We find evidence for an increased risk of infection by the Beta (B.1.351), Gamma (P.1), or Delta (B.1.617.2) variants compared to the Alpha (B.1.1.7) variant after vaccination. No clear differences were found between vaccines. However, the effect was larger in the first 14-59 days after complete vaccination compared to 60 days and longer. In contrast to vaccine-induced immunity, no increased risk for reinfection with Beta, Gamma or Delta variants relative to Alpha variant was found in individuals with infection-induced immunity.

## Introduction

Since the worldwide spread of severe acute respiratory syndrome coronavirus 2 (SARS-CoV-2) the virus has been slowly but steadily evolving. Although many nucleotide mutations are synonymous, multiple amino acid substitutions in functional domains of the spike protein are observed, some of which with likely impact on transmissibility, disease severity and pre-existing immunity^1^.

SARS-CoV-2 constellations of mutations under strong suspicion of a negative impact on virus epidemiology, virulence or effectiveness of social and public health measures (including diagnostics, vaccines, therapeutics) are designated Variant-of-Concern (VOC)^2^. As of 1 September 2021, four VOCs have been defined by the ECDC and WHO: Alpha (B.1.1.7, first detected in September 2020 in the United Kingdom), Beta (B.1.351, first detected in May 2020 in South Africa), Gamma (P.1, first detected in November 2020 in Brazil) and Delta (B.1.617.2, first detected in October 2020 in India)^2^. All four VOCs contain amino acid substitutions in the receptor binding domain (RBD) and N-terminal domain (NTD) of the Spike protein, which are known to be the main target of neutralizing antibodies. Several studies have shown decreased sensitivity of VOCs for convalescent and post-vaccination sera *in vitro*, with little to no reduction in sensitivity for the Alpha variant, the highest reduction in sensitivity for Beta and to a lesser extent for Gamma and Delta^3–6^.

These observations and the rapid global spread of first Alpha and later Delta sparked fear for SARS-CoV-2 escape from pre-existing immunity and selection of these variants in vaccinated and previously infected individuals. There are indications that the vaccine effectiveness (VE), especially against SARS-CoV-2 infection or mild COVID-19, is lower for the Beta, Gamma and Delta variant^7^. Less is known about the association between the observed VOCs and reinfection. Although an ecological study from the UK did not find an increase in the reinfection rate for the Alpha variant relative to pre-existing variants in the last quarter of 2020^8^, it needs to be determined if increased risk exists of reinfection by the Beta, Gamma, or Delta variants compared to the Alpha variant.

In January 2021, the COVID-19 vaccination program was rolled out in the Netherlands, first prioritizing health care workers, nursing home residents and the elderly. Current approved vaccines are either based on mRNA (Comirnaty, Spikevax) or on an Adeno-based vector system (Janssen COVID-19 vaccine, Vaxzevria) and are aimed to elicit a spike protein targeted humoral immune response that prevents virus entry and replication^9,10^. As of July 2021, all persons of 12 years and older have been offered COVID-19 vaccination. As of November 2021, 84% of all adults were fully vaccinated and 88% received at least one dose^11^. In the vaccination program in the Netherlands, Comirnaty (BNT162b2, BioNTech/Pfizer) has been used most often and has been offered to all age groups (76.0% of all administered doses). Spikevax (mRNA-1273, Moderna) has been mostly used in long term care facilities, health care workers, high medical risk groups and later also in the general population below 60 years (8.5% of all administered doses). Vaxzevria (ChAdOx1, AstraZeneca) has been used most in health care workers and the 60-65 years age group (12.1% of all administered doses). Janssen COVID-19 vaccine (Ad26.COV2.S, Janssen) has been used most in the 50-59 years age group and young adults (3.4% of all administered doses)^12^. Vaccination has proven to be highly effective against COVID-19, especially against hospitalization and death and reduces the secondary attack rate within households^7,13–17^.

Next to vaccination, infection with SARS-CoV-2 elicits a protective immune response though reinfections do occur. Studies comparing infection rates in the first and second surge of the SARS-CoV-2 pandemic between people who tested RT-PCR or antigen negative and positive in Denmark, Austria and Italy reported protection against repeat infection of 81%, 91% and 94%, respectively^18–20^. A prospective cohort study among health care workers in the UK found a 84% lower risk of infection after a previous infection^21^.

In the Netherlands, randomly selected SARS-CoV-2 RT-PCR positive specimens are sequenced to continuously monitor changes in the virus^22^. The Alpha variant started to increase rapidly from January 2021, and quickly became the dominant strain in the Netherlands. From June 2021, the Delta variant increased rapidly and caused nearly all infections from August 2021 onwards. In this study we aimed to investigate whether vaccine-or infection-induced immunity protects less well against infection by specific variants. Therefore, we compared the variant distribution of SARS-CoV-2 positive individuals who were either unvaccinated, vaccinated or had a previous infection using national epidemiological and molecular surveillance data from March up to August 2021.

## Methods

### Data

Persons testing positive for SARS-CoV-2 either by community testing or in a hospital are notified by Public Health Services (PHS) to the national surveillance database. Community testing is available through the PHS. Testing is encouraged in case of experiencing COVID-19-like symptoms, contact with a positive case, returning from another country, or upon a positive self-test. Data relevant for source and contact tracing and for surveillance is collected in the national surveillance database through a telephone interview, including data on vaccination status (i.e. number of doses, type of vaccine, and date of vaccination).

The Dutch national SARS-CoV-2 molecular surveillance program sequences whole virus genomes of randomly selected SARS-CoV-2 positive specimens from both community testing (via PHS) and hospitals, using a proper nationwide geographical distribution. In the current analysis, only samples with information on vaccination status and information on previous infection can be used. This information is collected in the national surveillance database and linked to sequence data using a sample identifier supplied during community testing. Sequences from hospital samples (5,893 out of the total 42,662 (13.8%) sequences of the SARS-CoV-2 genomic surveillance samples) and 7,464 of the 36,769 sequenced community samples were excluded as these could not be linked to the national surveillance database for required meta-data. In addition to randomly selected specimens, additional community testing specimens were requested for partially or fully vaccinated individuals as well as for cases with known prior laboratory-confirmed infection. This was done on a 3-weekly basis. This additional sampling resulted in an additional 1,516 cases to be included in the study and allowed for a detailed investigation of infecting variants after vaccination or reinfection. In the current analyses, cases with a sampling date between March 1 and August 31, 2021, were included.

### RT-PCR amplification and Nanopore sequencing

The majority of isolates were sequenced according to the following representative sequence method (minimal 85.3%), additional detailed protocols are available upon request. Total nucleic acid from combined nasopharyngeal and oropharyngeal swab was extracted using MagNApure 96 (MP96) with total nucleic acid kit small volume (Roche). Total nucleic acid was eluted in 50 μl Tris EDTA buffer.

SARS-CoV-2 specific RT-PCR amplification and sequencing was performed using the Nanopore protocol based on the ARTIC v3 amplicon sequencing protocol^23^. Several modifications to the protocol were made for optimization: 1) The total volume of the cDNA reaction is 12µl with a volume of 0.4µl Superscript IV instead of 0.6µl. 2) primer concentrations and primer sequence were adjusted for several amplicons to optimized amplicon yield and to match novel variants. Updated primer sequences are available upon request. 3) No distinction was made on the basis of Cp value, PCR was performed using 47 cycles. After the combination of PCR reactions A and B, the samples were quantified with the Qubit, samples with a concentration >35ng/μl were diluted to 6ng/µl in water. 5 µl of diluted PCR mix was used in the end-prep reaction. This end-prep is incubated for 15 min at 20°C and 15 min at 65°C. Barcoding was performed using the NEBNext Ultra II Ligation Module (E7595). In short, 1.3 µl end-prepped DNA was added to 2.5µl water, 6µl NEBNext Ultra II Ligation Master Mix, 0.2µl NEBNext Ligation Enhancer and 2 μl Native barcode SQK-LSK109 (EXP-NBD196). The Barcoding was incubated for 30 min at 20°C and 20 min at 65°C. Barcoded fragments were washed with twice with 870 µl short fragment buffer (SFB), once with 150 μl ethanol and eluted in 74 μl after 4 minute incubation with the beads. Adapter ligation was perfomed using NEBNext Quick Ligation Module (NEB) in a total volume 50 μl using 25 μl of AMPure XP beads. After washing with 125 μl short fragment buffer (SFB), the pellet was resuspended in 15.5 μl elution buffer. Finally, 45ng of library preparation was loaded on a flowcell (Nanopore) and sequencing was performed on a R9.4.1 flow cell multiplexing 48 up to 96 samples per sequence run for a run-time of 30 hours on a GridION (Nanopore).

GridION data was analyzed to get consensus genomes, with the SARS2seq pipeline and additional manual curation^24^. These genomes were analyzed with Pangolin (version 3.1.11) and NextClade (version 1.3.0) to get a final variant call^25,26^.

### Vaccination and previous infection status

Vaccination status is determined relative to the date used for statistics (DUFS). For symptomatic cases, this is the date of symptom onset or, if missing, the date of a positive test result minus 2 days. For asymptomatic cases, the DUFS is the date of positive test result. Fully vaccinated is defined as having received two doses of Comirnaty, Spikevax or Vaxzevria at least 14 days before DUFS or one dose of Janssen COVID-19 vaccine at least 28 days before DUFS. Partly vaccinated is defined as having received one dose of Comirnaty, Spikevax or Vaxzevria at least 14 days before DUFS, or two doses of Comirnaty, Spikevax or Vaxzevria less than 14 days before DUFS. A case is defined as recently vaccinated after one dose of Comirnaty, Spikevax or Vaxzevria 0-13 days or Janssen COVID-19 vaccine 0-27 days before DUFS. Individuals with a subsequent positive RT-PCR or antigen test result with an interval of at least 8 weeks after a previous positive test, including a period without symptoms, were defined as reinfections. This is either reported in the notification by the PHS or identified using record linkage by date of birth, sex, and 6-digit postal code.

### Statistical analyses

We compared the proportion of the four VOCs (Alpha, Beta, Gamma and Delta variant) between four immune status groups: 1) unvaccinated cases without a known previous infection (naïve), 2) partly vaccinated cases without a known previous infection, 3) fully vaccinated cases without a known previous infection, 4) unvaccinated cases with a previous infection. In a secondary analysis, fully vaccinated cases were further stratified by time between infection and last vaccination (<60 days versus >=60 days). Cases who were recently vaccinated, irrespective of their previous infection status, were excluded from the analyses, due to a possible incomplete immune response. Since the number of vaccinated cases with a previous infection was small (*n* = 111) this group was excluded.

The association between immune status and the Beta, Gamma and Delta variant was assessed using logistic regression. Immune status (group 2: partly vaccinated, group 3: fully vaccinated and group 4: previous infection versus group 1: naïve) was included in the model as the independent variable and Beta, Gamma or Delta vs Alpha as the dependent variable. We estimated odds ratios (ORs) with 95% confidence interval (CI) for any vaccine type and separately for Comirnaty, Spikevax, Vaxzevria and Janssen COVID-19 vaccine. An additional analysis was performed on the time since vaccination, stratifying the fully vaccinated by 14-59 and more than 60 days between complete vaccination and DUFS. As calendar time is both related to vaccination uptake and prevalence of a certain variant, i.e. a confounder, we included a natural cubic spline (5 knots) for calendar week of sample date in all regression models. In addition, all analyses were also adjusted for 10-year age group (40-49 years as reference) and sex.

## Results

From 1 March to 31 August 2021, a total of 661,658 SARS-CoV-2 positive cases were notified to the national surveillance database (Table 1). Of these, 38,261 (5.8%) cases were partly vaccinated, 25,933 (3.9%) were fully vaccinated and 10,565 (1.6%) had a known previous infection (Supplementary Figure 1). Of (partly) vaccinated cases, most received Comirnaty (65.0%), followed by Vaxzevria (19.3%), Janssen COVID-19 vaccine (9.8%) and Spikevax (5.9%). We included data of 29,305 samples that were sequenced through the national SARS-CoV-2 surveillance program (Table 1). In addition, 1,516 additional samples were sequenced to increase insight in variants present in infections after vaccination and reinfections were included.

**Table 1.** Characteristics of notified SARS-CoV-2 positive cases overall and for which variant information was available, 1 March to 31 August 2021, the Netherlands

|  | Notifications | Variant information from genomic surveillance | Variant information from additional sampling |
| --- | --- | --- | --- |
| Total | 661,658 | 29,305 | 1,516 |
| Immune status |  |  |  |
| Naïve | 487,063 (73.6%) | 20,804 (71.0%) | NA |
| Recently vaccinated | 47,565 (7.2%) | 2,140 (7.3%) | 18 (1.2%) |
| Partly vaccinated | 38,261 (5.8%) | 2,016 (6.9%) | 707 (46.6%) |
| Fully vaccinated | 25,933 (3.9%) | 1,791 (6.1%) | 516 (34.0%) |
| Previous infection | 10,565 (1.6%) | 284 (1.0%) | 191 (12.6%) |
| Vaccinated and previous infection | 2,065 (0.3%) | 62 (0.2%) | 49(3.2%) |
| Unknown | 50,206 (7.6%) | 2,208 (7.5%) | 35 (2.3%) |
| Age group |  |  |  |
| 0-9 | 42,666 (6.4%) | 1,818 (6.2%) | 4(0.3%) |
| 10-19 | 125,782 (19.0%) | 5,869 (20.0%) | 111 (7.3%) |
| 20-29 | 157,896 (23.9%) | 7,018 (23.9%) | 283 (18.7%) |
| 30-39 | 92,400 (14.0%) | 4,162 (14.2%) | 187 (12.3%) |
| 40-49 | 85,492 (12.9%) | 3,851 (13.1%) | 222 (14.6%) |
| 50-59 | 87,112 (13.2%) | 3,652 (12.5%) | 265 (17.5%) |
| 60-69 | 44,226 (6.7%) | 1,828 (6.2%) | 251 (16.6%) |
| 70-79 | 21,074 (3.2%) | 848 (2.9%) | 86 (5.7%) |
| 80+ | 5,010 (0.8%) | 259 (0.9%) | 107 (7.1%) |
| Sex |  |  |  |
| Male | 330,247 (49.9%) | 14,437 (49.3%) | 629 (41.5%) |
| Female | 331,411 (50.1%) | 14,868 (50.7%) | 692 (58.5%) |
| Symptoms |  |  |  |
| Yes | 556,214 (84.1%) | 25,478 (86.9%) | 1,355 (89.4%) |
| No | 66,593 (10.1%) | 2,248 (7.7%) | 121 (8.0%) |
| Unknown | 38,851 (5.9%) | 1,579 (5.4%) | 40 (2.6%) |
| Month (sampling date) |  |  |  |
| March | 149,103 (22.5%) | 5,408 (18.5%) | 177 (11.7%) |
| April | 171,534 (25.9%) | 4,621 (15.8%) | 335 (22.1%) |
| May | 114,536 (17.3%) | 4,874 (16.6%) | 137 (9.1%) |
| June | 24,904 (3.8%) | 3,162 (10.8%) | 97 (6.4%) |
| July | 146,978 (22.2%) | 6,620 (22.6%) | 438 (28.9%) |
| August | 54,603 (8.3%) | 4,620 (15.8%) | 331 (21.8%) |

Up to June 2021, 94.4% (14,068 of 14,903) of infections were caused by the Alpha variant, with a small proportion caused by the Beta (1.3%) and Gamma (1.3%) variant. The proportion of Delta increased from 0.9% (42 of 4874) in May to 98.7% (4561 of 4620) in August 2021. This pattern was observed over different immune statuses (Figure 1). In total, 17,890 (58.0%) Alpha, 209 (0.7%) Beta, 250 (0.8%) Gamma, 11,937 (38.7%) Delta and 535 (1.7%) other variant sequences were observed.

**Figure 1.**
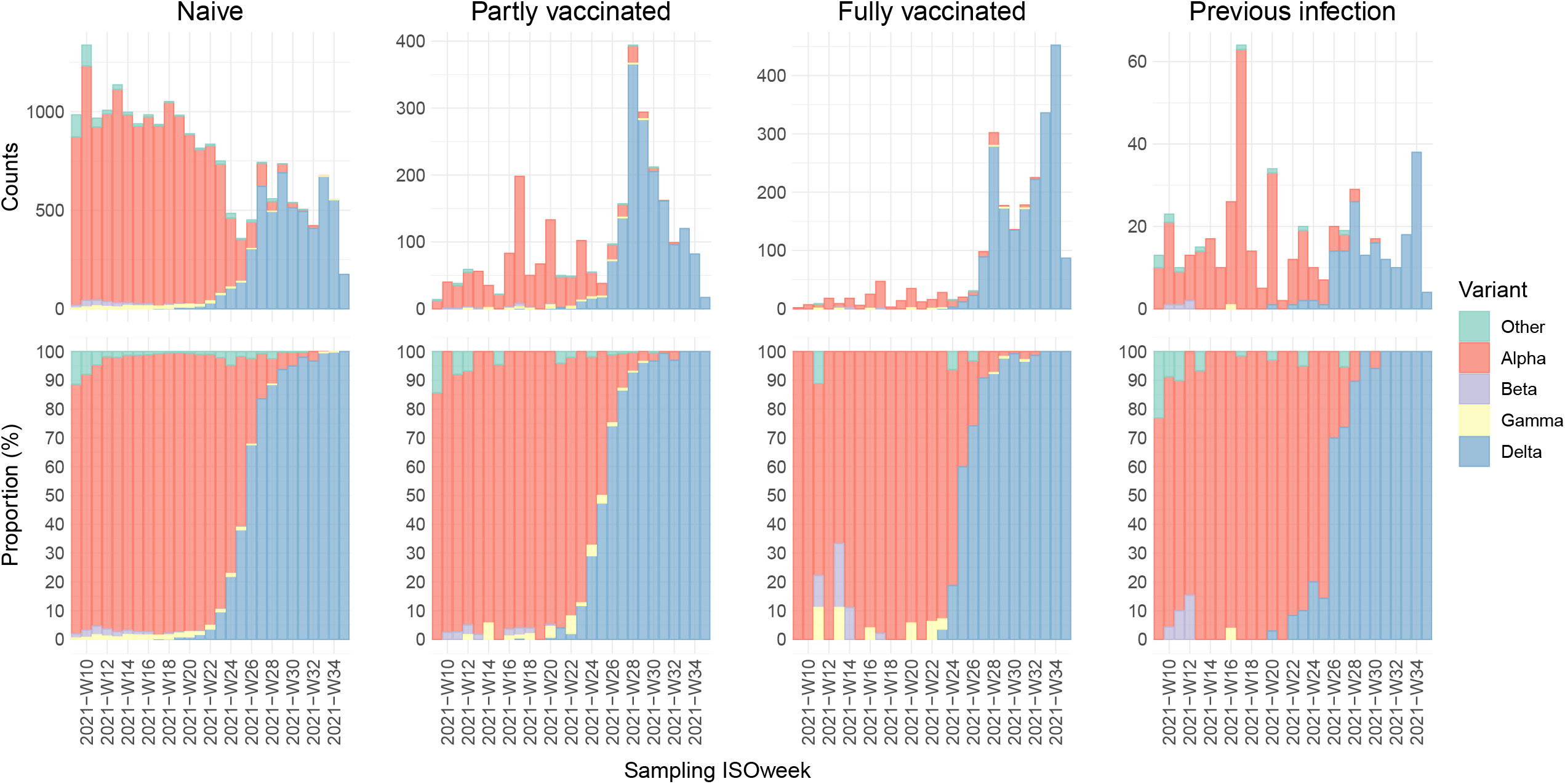
Variants found in SARS-CoV-2 positive samples of individuals with naïve (unvaccinated and no known previous infection), vaccine-induced, or infection-induced immune status. Number of naïve, partly vaccinated, fully vaccinated, and reinfected documented SARS-CoV-2 positive individuals by variant from March 1 to August 31, 2021 (upper panel) and proportion of the respective groups (lower panel).

Logistic regression analysis showed that full vaccination was significantly associated with infection with the Beta, Gamma or Delta variant compared to the Alpha variant (adjusted OR: 3.1 (95% CI: 1.3-7.3); 2.3 (95% CI: 1.2-4.4); 1.9 (95% CI: 1.4-2.5); respectively; Figure 2). The association for partial vaccination was less strong and not significant for Beta and Gamma, but still significant for Delta when compared to Alpha (adjusted OR: 1.6 (95% CI: 1.2-2.0); Figure 2). We did not find a significant association between previous infection and the Beta, Gamma or Delta variant over Alpha (adjusted OR: 1.4 (95% CI 0.5-3.7); 0.3 (95%CI 0.0-1.9; 1.0 (95%CI 0.6-1.5), respectively; Figure 2). The Delta variant was significantly associated with younger age groups, which highlights the importance of adjustment for age group (Supplementary Figure 3). When only including data from the genomic surveillance (excluding data from additional sampling of vaccinated and reinfected cases), similar odds ratios were found, although not significant anymore for Beta and Gamma due to less power (data not shown).

**Figure 2.**
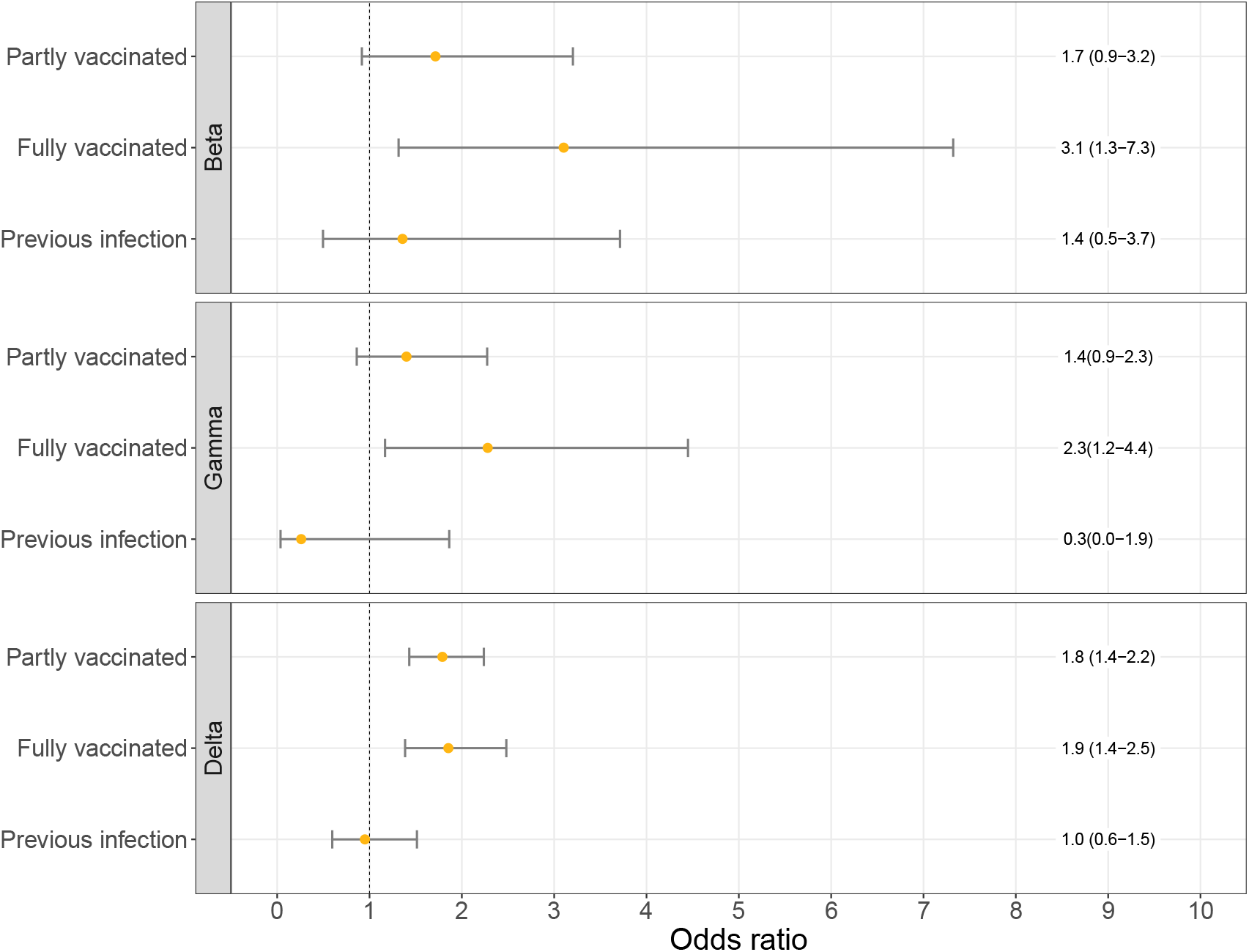
Odds ratios of the logistic regression models for the association between immune status and VOC (Beta, Gamma or Delta over the Alpha variant) adjusted for week of sampling, sex and 10-year age group. Error bars correspond to the 95% confidence intervals.

When stratified by vaccine type, the association between full vaccination and infection with the Delta variant was significant for Comirnaty and Janssen COVID-19 vaccine, but not for Spikevax and Vaxzevria (Table 2). The association between partial vaccination and the Delta variant was significant for Comirnaty and Vaxzevria but not Spikevax. In addition, we stratified the fully vaccinated by time since vaccination. The association for individuals with less time (14-59 days) between onset and last dose was higher (OR: 2.3 (95%CI 1.6-3.4)) compared to individuals with 60 days and more (OR: 1.4 (95%CI 1.0-2.1)) for the Delta variant. A similar trend was observed for Beta variant and Gamma variant, although with wide confidence intervals (Table 2).

**Table 2.** Odds ratios (ORs) and 95% confidence intervals (CIs) for the association between immune status and VOC (Beta, Gamma or Delta over the Alpha variant) by vaccine type and days between onset and last dose, both adjusted for week of sampling, sex and 10-year age group.

|  | Beta<br>OR (95% CI) | Gamma<br>OR (95% CI) | Delta<br>OR (95% CI) |
| --- | --- | --- | --- |
| Naïve | Reference | Reference | Reference |
| Partly vaccinated |  |  |  |
| Comirnaty | 1.1 (0.3-3.7) | 1.9 (1.0-3.7) | 1.8 (1.4-2.3) |
| Spikevax | n/a | 2.5 (0.6-10.4) | 1.1 (0.6-2.0) |
| Vaxzevria | 2.1 (1.0-4.1) | 1.0 (0.5-2.0) | 2.1 (1.3-3.5) |
| Fully vaccinated |  |  |  |
| Comirnaty | 3.2 (1.4-7.7) | 2.2 (1.0-4.7) | 2.2 (1.4-3.3) |
| Spikevax | n/a | n/a | 1.3 (0.4-4.3) |
| Vaxzevria | n/a | 2.8 (0.7-12.3) | 1.4 (0.9-2.3) |
| Janssen | n/a | 4.4 (0.6-34.5) | 2.2 (1.2-4.2) |
| Naïve | Reference | Reference | Reference |
| Fully vaccinated |  |  |  |
| 14-60 days | 3.7 (1.4-9.5) | 3.0 (1.3-7.1) | 2.3 (1.6-3.4) |
| >60 days | 1.7 (0.2-12.8) | 1.7 (0.6-4.6) | 1.4 (1.0-2.1) |

## Discussion

Using national epidemiological and whole genome sequencing surveillance data from March to August 2021 in the Netherlands, our analysis provides evidence for an increased risk of infection by the Beta, Gamma, or Delta variants compared to the Alpha variant after full vaccination, regardless of the vaccine used. This indicates lower vaccine effectiveness against infection with the Beta, Gamma and Delta variant compared to the Alpha variant. No clear differences between vaccine type were observed as confidence intervals largely overlap. Interestingly, we did not find a significant difference between susceptibility to any of the investigated VOCs among individuals with immunity due to a previous infection compared to naïve individuals. Also when stratified by time between infections no differences are observed (data not shown). Of note is that these analyses do not aim to determine the probability of getting infected after vaccination or previous infection, but rather calculate the likelihood of getting infected with specific VOCs.

The association with vaccination status was higher for Beta and Gamma (OR of 3.1 and 2.3, respectively) than for Delta (OR of 1.9), although confidence intervals for Beta and Gamma were wide because of low numbers. This is in line with literature showing lower vaccine effectiveness estimates against infection for Beta and Gamma compared to Delta^7^. An OR for Delta of 1.9 implicates a reduction of vaccine effectiveness from 90% to 80%, which has been shown in the UK^3,27^. Current literature still shows high vaccine effectiveness of 90-95% against severe COVID-19 for the Delta variant^7,17^, which is reassuring. However, note that with very high vaccine effectiveness, a difference of a factor 1.5-2.0 between two variants could go unnoticed, as it would only mean a decrease of effectiveness of 95 to 92%.

Spike binding and neutralization have been shown to be substantially reduced against Beta, Gamma, and Delta with the largest reduction in neutralization against Beta^4–6^, which is consistent with our results. This observation did not differ for infection- or vaccine-induced immunity, although convalescent sera from mild infections showed lower levels of neutralization potency to VOCs compared to hospitalized cases and vaccinated individuals^4^. However, in Alpha and Beta a reduction was not observed for T-cell-mediated immunity^28^.

We observed a larger effect of vaccination in the first 14-59 days after vaccination (i.e. OR 2.3 (95%CI 1.6-3.5) for Delta) compared to 60 days and longer (i.e. OR 1.4 (95%CI 1.0-2.1) for Delta), suggesting that the difference in VE between Delta and Alpha variant reduces over time since vaccination, possibly due to waning immunity. A recent large cohort study describes an effect of waning and a small effect of the circulating variant (i.e. Delta vs non-Delta) on the VE against SARS-CoV-2 infection^29^. They find a non-delta VE of 97% and a delta VE of 93% one month after vaccination, which means a ratio of 2.3 between non-delta VE and delta VE. Four to five months post vaccination, VE estimates of 67% and 53% for non-delta and delta were observed respectively, a ratio of 1.4. This very well corresponds with our results. Given the broad and sometimes overlapping confidence intervals of these data, however, the differences need to be interpreted with caution.

We found no association between previous infection and a new infection with Beta, Gamma or Delta versus Alpha, suggesting that there is a no difference in immunity between Alpha and Beta, Gamma or Delta after previous infection, in contrast to vaccine-induced immunity. It is not yet clear whether previous infection or vaccination induces better protection against infection. However, primary infection comes with a risk of hospitalization or death, especially in older persons or individuals with underlying conditions. Even if infection-induced immunity protects better against reinfection with novel variants, vaccination is preferred over infection to protect individuals against severe disease as the cumulative risk from two infections should be taken into account.

Some limitations of our study need to be addressed. Asymptomatic or mild cases with low viral load are less likely to be identified and only detectable infections can be sequenced and included. In addition, sequencing is more successful in samples with low to medium Ct values (high to medium viral load). If infection with Beta, Gamma or Delta leads to lower Ct values than Alpha and Ct values are higher for infections after vaccination^30–32^, this could have led to an overestimation of the studied association. Another limitation is that prior infections could go undetected, especially if occurred during the first wave when there was no mass scale testing capacity. This could lead to an underestimation of cases with a previous infection, as we do not directly measure pre-existing infection-induced immunity.

In conclusion, our results confirm a lower vaccine effectiveness against infection for the Delta variant, and similarly the Beta and Gamma variant, compared to Alpha. This effect was largest early after complete vaccination. These findings are informative for considerations on vaccine updates, future vaccination and pandemic control strategies.

## Data Availability

Data produced in the present study are available upon reasonable request to the authors.

## Acknowledgements

The authors would like to thank all personnel at the 25 Public Health Services for data collection in the national surveillance database and all laboratories for providing specimens for sequence analyses.

## Code availability

Code for sequencing data processing is publicly available at github.com/RIVM-bioinformatics/SARS2seq. Scripts for statistical analysis, figures, and tables can be found at github.com/Stijn-A/xxxxx.[upon publication]

## References

1. Oude Munnink, B. B. et al. The next phase of SARS-CoV-2 surveillance: real-time molecular epidemiology. Nat. Med. 2021 279 27, 1518–1524 (2021).

2. WHO. Tracking SARS-CoV-2 variants. https://www.who.int/en/activities/tracking-SARS-CoV-2-variants/ (2021).

3. Bernal, J. L. et al. Effectiveness of Covid-19 Vaccines against the B.1.617.2 (Delta) Variant. https://doi.org/10.1056/NEJMoa2108891 385, 585–594 (2021).

4. Caniels, T. G. et al. Emerging SARS-CoV-2 variants of concern evade humoral immune responses from infection and vaccination. Sci. Adv. 7, (2021).

5. Bates, T. A. et al. Neutralization of SARS-CoV-2 variants by convalescent and BNT162b2 vaccinated serum. Nat. Commun. 2021 121 12, 1–7 (2021).

6. Liu, J. et al. BNT162b2-elicited neutralization of B.1.617 and other SARS-CoV-2 variants. Nat. 2021 5967871 596, 273–275 (2021).

7. Krause, P. R. et al. Considerations in boosting COVID-19 vaccine immune responses. Lancet 0, (2021).

8. Graham, M. S. et al. Changes in symptomatology, reinfection, and transmissibility associated with the SARS-CoV-2 variant B.1.1.7: an ecological study. Lancet Public Heal. 6, e335–e345 (2021).

9. Dai, L. & Gao, G. F. Viral targets for vaccines against COVID-19. Nat. Rev. Immunol. 2020 212 21, 73–82 (2020).

10. van Gils, M. J. et al. Four SARS-CoV-2 vaccines induce quantitatively different antibody responses against SARS-CoV-2 variants Short title: Vaccine antibody responses against SARS-CoV-2 variants Amsterdam UMC COVID-19 S3/HCW study group. doi:10.1101/2021.09.27.21264163.

11. RIVM. Deelname COVID-19-vaccinatie in Nederland. https://www.rivm.nl/sites/default/files/2021-11/COVID-19_Vaccinatie_Schattingen_WebSite_rapport_20211101_1552_def.pdf (2021).

12. VWS. COVID-19 vaccinations. coronadashboard.government.nl/landelijk/vaccinaties (2021).

13. Higdon, M. M. et al. A systematic review of COVID-19 vaccine efficacy and effectiveness against SARS-CoV-2 infection and disease. medRxiv 2021.09.17.21263549 (2021) doi:10.1101/2021.09.17.21263549.

14. Harder, T. et al. Efficacy and effectiveness of COVID-19 vaccines against SARS-CoV-2 infection: interim results of a living systematic review, 1 January to 14 May 2021. Eurosurveillance 26, 2100563 (2021).

15. Gier, B. de et al. Vaccine effectiveness against SARS-CoV-2 transmission and infections among household and other close contacts of confirmed cases, the Netherlands, February to May 2021. Eurosurveillance 26, 2100640 (2021).

16. Gier, B. de et al. Vaccine effectiveness against SARS-CoV-2 transmission to household contacts during dominance of Delta variant (B.1.617.2), the Netherlands, August to September 2021. Eurosurveillance 26, 2100977 (2021).

17. Gier, B. de et al. COVID-19 vaccine effectiveness against hospitalizations and ICU admissions in the Netherlands, April-August 2021. medRxiv 2021.09.15.21263613 (2021) doi:10.1101/2021.09.15.21263613.

18. Hansen, C. H., Michlmayr, D., Gubbels, S. M., Mølbak, K. & Ethelberg, S. Assessment of protection against reinfection with SARS-CoV-2 among 4 million PCR-tested individuals in Denmark in 2020: a population-level observational study. Lancet 397, 1204–1212 (2021).

19. Pilz, S. et al. SARS-CoV-2 re-infection risk in Austria. Eur. J. Clin. Invest. 51, e13520 (2021).

20. Leidi, A. et al. Risk of Reinfection After Seroconversion to Severe Acute Respiratory Syndrome Coronavirus 2 (SARS-CoV-2): A Population-based Propensity-score Matched Cohort Study. Clin. Infect. Dis. (2021) doi:10.1093/CID/CIAB495.

21. Hall, V. J. et al. SARS-CoV-2 infection rates of antibody-positive compared with antibody-negative health-care workers in England: a large, multicentre, prospective cohort study (SIREN). Lancet 397, 1459–1469 (2021).

22. Variants of the coronavirus SARS-CoV-2 | RIVM. https://www.rivm.nl/en/coronavirus-covid-19/virus-sars-cov-2/variants.

23. nCoV-2019 sequencing protocol v2 (GunIt). https://www.protocols.io/view/ncov-2019-sequencing-protocol-v2-bdp7i5rnãversion_warning=no.

24. GitHub -RIVM-bioinformatics/SARS2seq: SARS2seq is a pipeline designed to process raw FastQ data from targeted SARS-CoV-2 sequencing and generate biologically correct consensus sequences of the SARS-CoV-2 genome. https://github.com/RIVM-bioinformatics/SARS2seq.

25. Aksamentov, I., Roemer, C., Hodcroft, E. B. & Neher, R. A. Nextclade: clade assignment, mutation calling and quality control for viral genomes. (2021) doi:10.5281/ZENODO.5607694.

26. Rambaut, A. et al. A dynamic nomenclature proposal for SARS-CoV-2 lineages to assist genomic epidemiology. Nat. Microbiol. 2020 511 5, 1403–1407 (2020).

27. Sheikh, A., McMenamin, J., Taylor, B. & Robertson, C. SARS-CoV-2 Delta VOC in Scotland: demographics, risk of hospital admission, and vaccine effectiveness. Lancet 397, 2461–2462 (2021).

28. Geers, D. et al. SARS-CoV-2 variants of concern partially escape humoral but not T-cell responses in COVID-19 convalescent donors and vaccinees. Sci. Immunol. 6, 1750 (2021).

29. Tartof, S. Y. et al. Effectiveness of mRNA BNT162b2 COVID-19 vaccine up to 6 months in a large integrated health system in the USA: a retrospective cohort study. Lancet 0, (2021).

30. Singanayagam, A. et al. Community transmission and viral load kinetics of the SARS-CoV-2 delta (B.1.617.2) variant in vaccinated and unvaccinated individuals in the UK: a prospective, longitudinal, cohort study. Lancet Infect. Dis. 0, (2021).

31. Levine-Tiefenbrun, M. et al. Viral loads of Delta-variant SARS-CoV-2 breakthrough infections after vaccination and booster with BNT162b2. Nat. Med. 2021 1–3 (2021) doi:10.1038/s41591-021-01575-4.

32. Luo, C. H. et al. Infection with the SARS-CoV-2 Delta Variant is Associated with Higher Infectious Virus Loads Compared to the Alpha Variant in both Unvaccinated and Vaccinated Individuals. medRxiv 2021.08.15.21262077 (2021) doi:10.1101/2021.08.15.21262077.

