## Supplementary information for "Increased risk of infection with SARS-CoV-2 Beta, Gamma, and Delta variant compared to Alpha variant in vaccinated individuals"

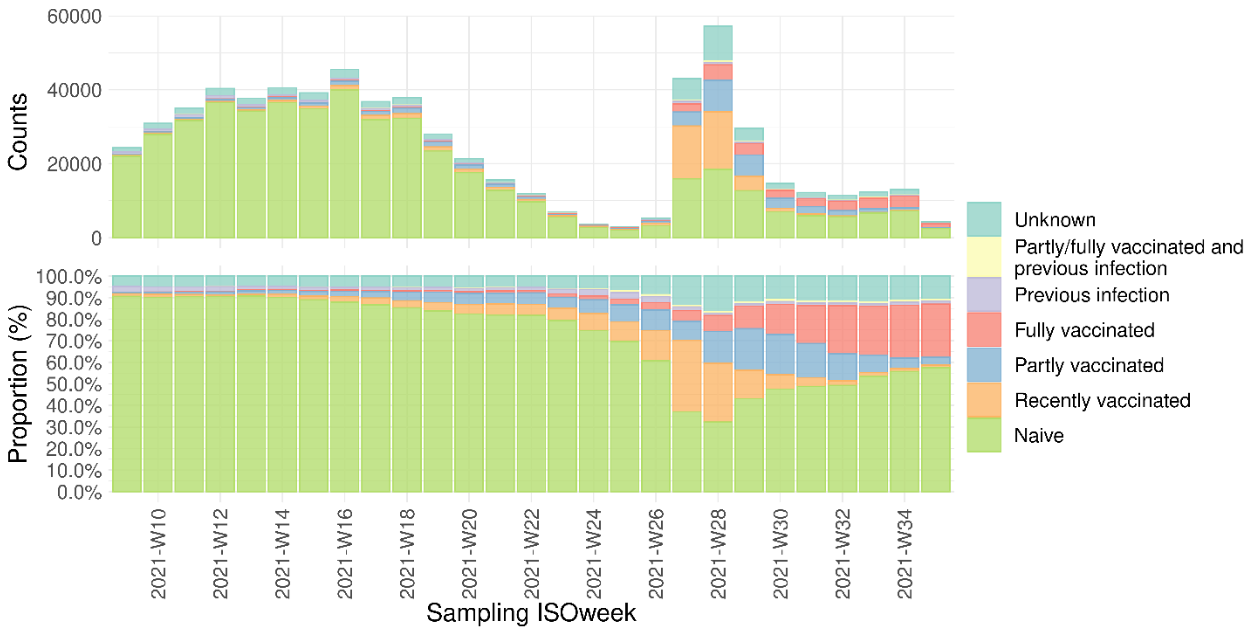


***Supplementary Figure 1*** *Immune status of notified SARS-CoV-2 cases in the Netherlands. Number of naïve (unvaccinated and no known previous infection), recently vaccinated, partly vaccinated, fully vaccinated, reinfected, partly/fully vaccinated and reinfected, and unknown documented SARS-CoV-2 positive individuals from March 1 to August 31, 2021 (upper panel) and proportion of the respective groups (lower panel).*

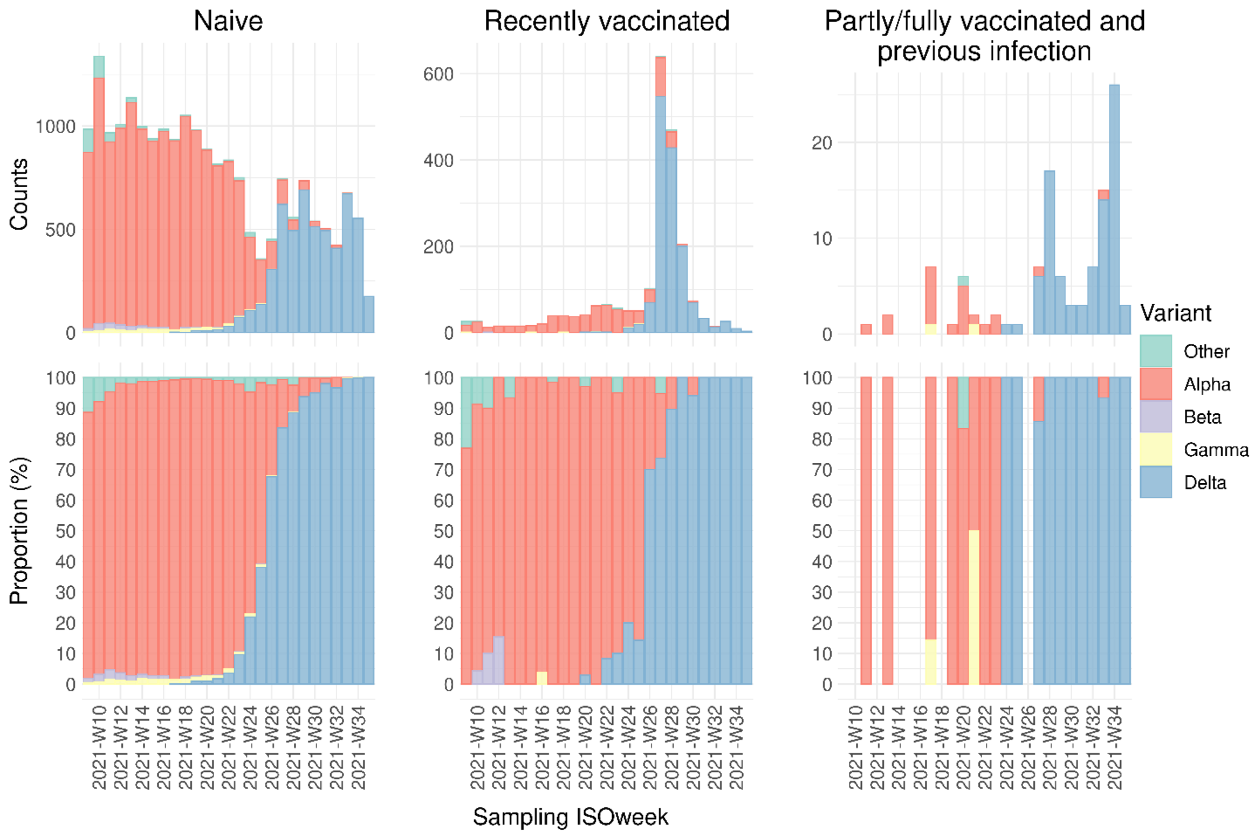


***Supplementary Figure 2*** *Variants found in SARS-CoV-2 positive samples of individuals with naïve (unvaccinated and no known previous infection), recently vaccinated, or vaccine-induced and infection-induced immune status. Number of naïve, recently vaccinated, partly/fully vaccinated and reinfected documented SARS-CoV-2 positive individuals by variant from March 1 to August 31, 2021 (upper panel) and proportion of the respective groups (lower panel).*

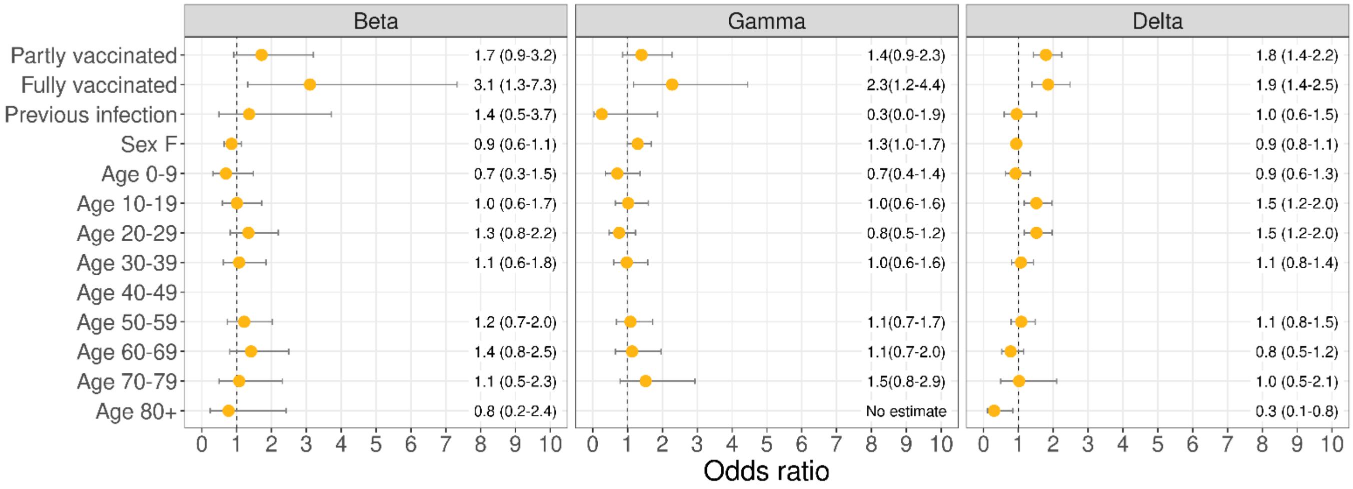


***Supplementary Figure 3*** *Odds ratios and 95% confidence intervals for the association between immune status and the Beta, Gamma or Delta variant with adjustment for week of sampling, 10-year age group (40-49 is reference) and sex.*
